# Mapping Respiratory Health Digital Interventions in South and Southeast Asia: A Scoping Review

**DOI:** 10.1101/2024.12.12.24318897

**Authors:** Laura Evans, Jay Evans, Adina Abdullah, Zakiuddin Ahmed, The RESPIRE Group

## Abstract

**Background:** Digital health has progressed rapidly due to the advances in technology and the promises of improved health and personal health empowerment. Concurrently, the burden of respiratory disease is increasing, particularly in Asia, where mortality rates are higher, and public awareness and government engagement are lower than in other regions of the world. Leveraging digital health interventions to manage and mitigate respiratory disease presents itself as a potentially effective approach. This study aims to undertake a scoping review to map respiratory digital health interventions in South and Southeast Asia, identify existing technologies, opportunities, and gaps, and put forward pertinent recommendations from the insights gained.

**Methods:** This study used a scoping review methodology as outlined by Arksey and O’Malley and the Joanna Briggs Institute. Medline, Embase, CINAHL, PsycINFO, Cochrane Library, Web of Science, PakMediNet and MyMedR databases were searched along with key websites grey literature databases.

**Results:** This scoping review has extracted and analysed data from 87 studies conducted in 14 South and Southeast Asian countries. Results were mapped to the WHO classification of digital health interventions categories to better understand their use. Digital health interventions are primarily being used for communication with patientes and between patients and providers. Moreover, interventions targeting tuberculosis were the most numerous. Many ‘old’ interventions, such as SMS, are still being used but updated. Artificial intelligence and machine learning are also widely used in the region at a small scale. There was a high prevalence of pilot interventions compared to mature ones.

**Conclusions:** This scoping review collates and synthesises information and knowledge in the current state of digital health interventions, showing that there is a need to evaluate whether a pilot project is needed before starting, there is a need to report on interventions systematically to aid evaluation and lessons learnt, and that artificial intelligence and machine learning interventions are promising but should adhere to best ethical and equity practices.

**Author summary:** Technology has advanced quickly, facilitating the development of digital health, that is the use of technological tools for health purposes. Digital health tools may help more people achieve better health. At the same time, respiratory diseases are becoming a growing problem, especially in Asia, where there are more deaths and diseases linked to respiratory causes than in other parts of the world. Using digital health tools may be an effective way to manage and reduce the impact of respiratory diseases in the region. To that end, this study reviewed current digital health tools in South and Southeast Asia, identified gaps and opportunities and made recommendations based on the findings. The methodology used was a scoping review, which followed standards as described by Arksey and O’Malley and the Joanna Briggs Institute. It searched relevant medical databases for information. This review includes 87 studies from 14 different countries. It revealed that tuberculosis was the most targeted disease by digital health interventions and that older technologies, such as the SMS, are still being used and updated as needed. Moreover, it revealed that new technologies like artificial intelligence and machine learning are being used more frequently but in small projects and that many of the projects described are small-scale pilot projects.

## Introduction

Digital health, that is, the use of information communication technologies to manage illnesses and wellness, has, in the last two decades, progressed rapidly due to the advances in technology and the promises of improved health and personal health empowerment (1,2). Concurrently, the burden of respiratory disease is increasing, with both infectious and non-communicable respiratory diseases contributing significantly to years of life lost due to premature death and years lived with a disability (3,4). This increased burden of respiratory disease is a significant reality in Asia, where mortality rates are higher, and public awareness and government engagement are lower than in other regions of the world (5,6). Leveraging digital health interventions (DHIs) to manage and mitigate respiratory disease and its adverse health effects presents itself as a potentially effective approach. However, a critical step to harnessing the power of digital health must be understanding the current digital landscape. Knowing what current DHIs are being deployed and used successfully (or not) may help reduce costs, avoid duplication, and increase the efficiency, accessibility, and sustainability of such interventions (7–10).

The Global Health Research Unit on Respiratory Health (RESPIRE) is a collaboration between several Asian organisations and universities in Bangladesh, Bhutan, India, Malaysia, Pakistan, Indonesia, and Sri Lanka and the University of Edinburgh in Scotland, UK (6,11,12). RESPIRE aims to ‘develop into a world-leading unit that will: (i) map and collate continuing and emerging respiratory challenges; (ii) prioritise existing evidence-based interventions that have the potential to be adapted to reduce mortality and morbidity in low- and middle-income countries (LMICs); (iii) support local adaptation and tailoring of interventions for deployment in low-resource environments and catalyse developmental work in areas of unmet need; (iv) support local implementation efforts and evaluation of programmes of work; and (v) identify the best delivery mechanisms for long-term delivery and scaling-up’. (6) RESPIRE partners have a long-standing track record of developing and deploying DHIs in challenging settings such as digital auscultation, computational frameworks to interpret chest x-rays, home-based pulmonary rehabilitation, mobile-based support for Community Health Workers, remote teleconsultation, mobile-based interventions for adults with asthma and blended learning for professionals (7). In order to enhance and support the wide range of digital health initiatives of RESPIRE partners in South and Southeast Asia, as well as other stakeholders, our goal was to create a landscape mapping assessment of the current state of DHIs in the region.

Understanding the current landscape of digital health interventions that target respiratory diseases will (i) uncover existing gaps, (ii) highlight potential opportunities, (iii) suggest research and programme priorities most needed in the field of digital health to address current respiratory health diseases in South and Southeast Asia, and (iv) further advance RESPIRE’s overall aims. Therefore, we aim to undertake a scoping review to map respiratory digital health interventions in South and Southeast Asia, identify existing technologies, opportunities, and gaps, and put forward pertinent recommendations from the insights gained.

## Methods

### Scoping review methodology

This study used a scoping review methodology as outlined by Arksey and O’Malley and the Joanna Briggs Institute (JBI) (13,14). Specific details of the methods for this study can be found in the previously published protocol (15). The scoping review methodology is an appropriate approach for mapping digital health interventions because it allows flexibility when exploring the diverse and heterogenous field of digital health, is suitable when using different sources of data (peer-reviewed, grey, expert opinions, etc.), and permits inclusion and exclusion criteria to be iteratively refined as more evidence is uncovered (13,14,16,17). Additionally, the PRISMA extension for scoping reviews (PRISMA-ScR) (18) will be followed to ensure adherence to methodological standards (this can be referenced in the supplementary materials).

### Identifying the research question

This scoping review sought to answer the following research questions:

1. What digital health tools and technologies are being employed in South and Southeast Asia for respiratory health?
2. How are these addressing (or not) the respiratory health needs of the region?
3. What recommendations can be made from the literature?

### Identifying relevant studies

In order to identify relevant literature, the Medline, Embase, CINAHL, PsycINFO, Cochrane Library, Web of Science, PakMediNet and MyMedR databases were searched. Additionally along with the following key websites grey literature databases: ProQuest Thesis and Dissertations, Digital Health Atlas, Global Digtial Health Monitor, World Health Organisation’s Global Index Medicus.

The search strategy was initially drafted for Medline in the Ovid platform (Table 2) and was adapted for the remaining databases. The search strategy was iteratively developed and refined by the authors’ input and the librarian at the University of Edinburgh. The terms ‘respiratory health’, ‘digital health’, ‘South Asia’, ‘Southeast Asia’, and all relevant variations of these terms were included in the search strategy to gather as much pertinent literature and information as possible.

**Table 1.** Search strategy.

|  |  |
| --- | --- |
| **1** | occupational diseases/ or Asthma/ or Air Pollution/ or Respiratory Tract Diseases/ or Occupational Exposure/ or Air Pollutants/ or Tuberculosis/ or Tuberculosis, Pulmonary/ or "Tobacco Use"/ or Tobacco/ or "Tobacco Use Cessation"/ or "Tobacco Use Cessation Devices"/ or Pulmonary Disease, Chronic Obstructive/ or Pneumonia/ or COVID-19/ |
| **2** | (respiratory health or tuberculosis or tobacco or pneumonia).mp |
| **3** | 1 or 2 |
| **4** | telemedicine/ or telehealth/ or artificial intelligence/ or machine learning/ or medical informatics/ or electronic health records/ or mobile applications/ or exp Informatics/ |
| **5** | (artificial intelligence or digital health or e-health or ehealth or m-health or mhealth or (digital adj2 (health or solution* or system*)) or (health adj2 (electronic or record* or tele*)) or ict4d or (information adj5 development) or machine learning or mobile health or telecare or telehealth or telemedicine or tele-health or teleconsultation or tele-consultation or tele-care or tele-medicine or (tele adj1 (medicine or care or health or consultation)) or ((virtual* or remote*) adj4 (visit* or consult* or meet* or appoint* or communicat*)) or (Health* adj4 tech*) or e-portal* or eportal* or (Patient* adj2 portal*) |
|  | or (medical adj2 informatic*).mp. |
| **6** | 4 or 5 |
| **7** | Asia/ or Asia, Southern/ or Asia, Southeastern/ |
| **8** | (Asia or Brunei Darussalam or Cambodia or Indonesia or Lao People's Democratic Republic or Malaysia or Myanmar or Philippines or Singapore or Thailand or Timor-Leste or Viet Nam or Vietnam or Afghanistan or Bangladesh or Bhutan or India or Iran or Islamic Republic of Iran or Maldives or Nepal or Pakistan or Sri Lanka).mp |
| **9** | 7 or 8 |
| **10** | 3 and 6 and 9 |

**Table 2.** Inclusion and exclusion criteria.

| **Domain** | **Inclusion criteria** | **Exclusion criteria** |
| --- | --- | --- |
| Population | Any population | N/A |
| Concept | Technological interventions for respiratory health that fall | Other non-technological interventions used for respiratory health |
|  | under any of the categories of the World Health Organization's (WHO) classification of digital health interventions.<br>Respiratory health includes respiratory infections, non-communicable respiratory diseases, and preventable risk factors for respiratory conditions as defined by RESPIRE | Not a respiratory health focus. |
| Context | South and Southeast Asia | Rest of the world |
| Types of evidence sources | Any study design | Books<br>Abstracts only<br>Posters<br>Protocols |
| Other variables | Published in English<br>Studies or data published in the last ten years (2013-2023)<br>Full article/data available digitally | Published in any other language<br>Studies or data published before 2013<br>Full article/data not available digitally |

### Study selection

Exclusion and inclusion criteria were developed according to the domains suggested by the JBI (16,18) of population, concept, context, and type of evidence. Additionally, an ‘other variables’ category was created to include year of pubication, language, and format criteria (table 3). The regions of South and Southeast Asia include 19 countries as established by the United Nations Statistics Division (19). These two regions have been chosen because they contain all the RESPIRE2 countries. Multi-country studies containing countries from the selected regions and other regions of the world were included in the initial screening and only excluded if they did not provide the data of interest separately for each country. Only studies in English and those published since 2013 were included to keep the scope of this review within manageable boundaries.

For information found in a scientific study format, Covidence software (20) was used to eliminate duplicates and perform screening. After deduplication, title, abstract, and full-text screening will be done by two authors according to the inclusion and exclusion criteria. Discrepancies were first addressed by consensus between those two authors. If there was a lack of consensus, a third reviewer decided.

For all other types of information or data found from searches, manual screening by one reviewer happened first. Relevant data was entered into a spreadsheet, and a second reviewer assessed it. Discrepancies were first addressed between both reviewers, and if there was no consensus, a third reviewer made the final decision. We did not contact authors directly to further understand whether a study should be included or not since it would most likely lengthen the timeline for this scoping review.

Database searches yielded 11162 studies that were imported into Covidence. Of those, 687 were duplicates and removed. 10465 study titles were screened, and 10285 were found irrelevant and excluded. The abstract and full-text screening was done in 180 studies; 94 were excluded for various reasons (see PRISMA ScR table below, figure 1), and 86 were included for analysis. Of the studies included for analysis, three were studies with more than one publication in the same study. Those were combined as one study for analysis purposes. The studies included were not assessed for quality beyond the inclusion criteria as per scoping review guidelines (16).

**Figure 1:**
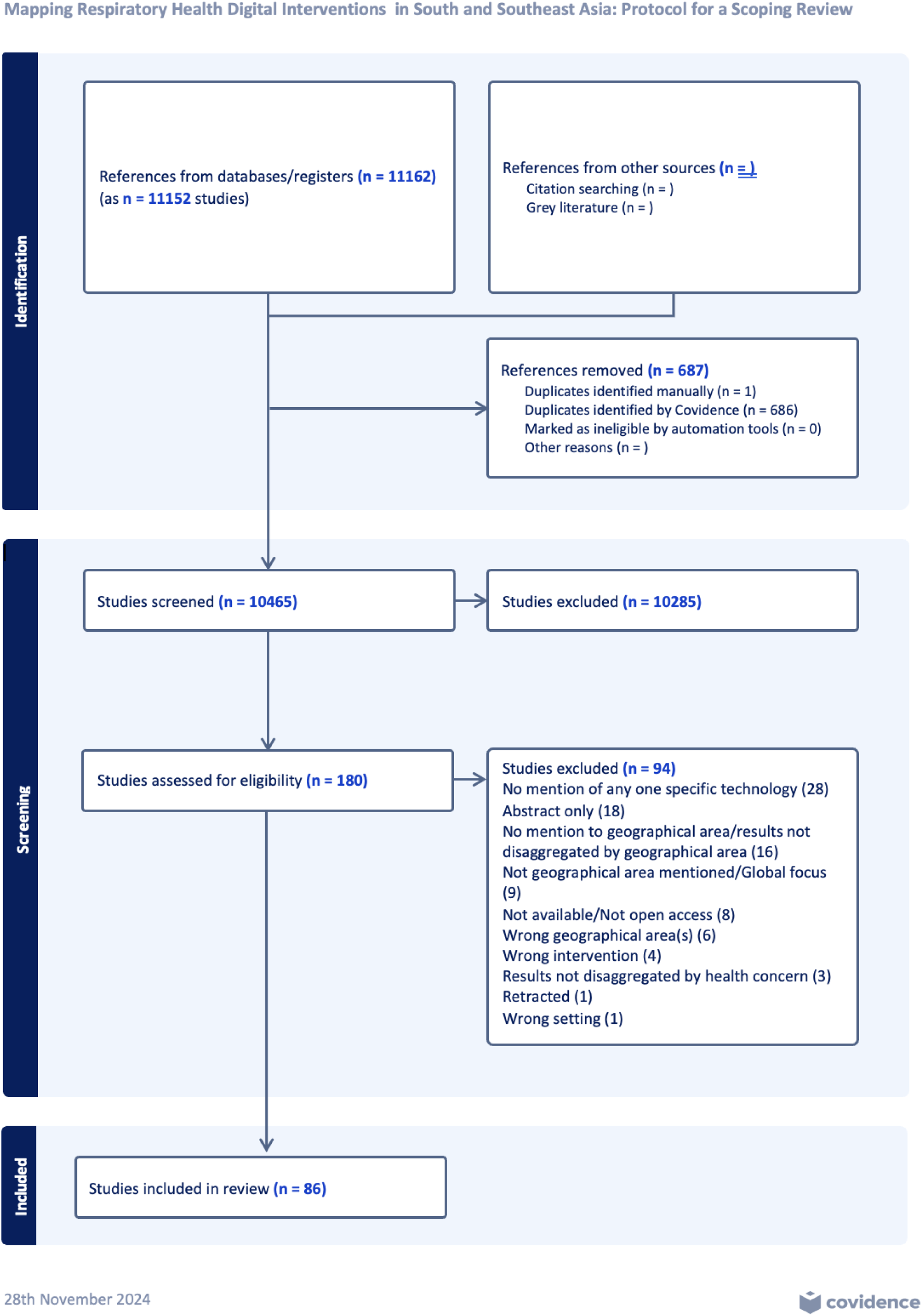
PRISMA ScR for this study (18)

### Ethical considerations

Scoping reviews use secondary data and do not require ethics approval under RESPIRE rules. However, the authors adhered to the highest ethical standards when carrying out the review. This methodology establishes a transparent and reproducible study design, which limits the potential for personal bias (21).

Generative artificial intelligence (AI) was not used at any time during the research and writing of this scoping review. However, Grammarly (22), a tool for checking grammatical correctness that uses AI in the background, was employed during the writing of this manuscript.

### Charting the data

After selection, relevant data was extracted to a spreadsheet. The extraction form was first piloted in 3-5 studies to assess if it was fit for purpose as recommended by the JBI. After this pilot, several columns were added: intervention (listing the name of the intervention if it had one or if not the type), results (listing the results of the intervention according to the authors), target disease (listing their intended target disease), pilot (a yes or no column to identify which interventions were pilot studies and which ones were beyond the pilot study phase). Also, one of the columns was divided into two: the type (name) of intervention according to the WHO’s classification of digital health interventions and the code of the intervention according to the same classification to facilitate analysis later on.

Data from a non-scientific study format was very limited because although the grey literature databases searched contained several interventions, most times, the information available was insufficient to evaluate their inclusion in the study according to the established criteria.

### Consultation

Consultation with stakeholders was ongoing throughout the scoping review process. We disseminated early findings among partners via a poster presentation, allowing them to provide feedback on findings to be incorporated into the discussion.

## Results

The supplementary material in the extraction spreadsheet used for this review contains full results from the included studies. Below are the most relevant results for our research questions.

### Countries represented

This scoping review has extracted and analysed data from 87 studies conducted in 14 South and Southeast Asian countries (Figure 2). India had the highest number of studies at 34, followed by Indonesia with 12 and Pakistan with 10. Three studies involved two or more countries. Complete numbers for each country can be seen in the legend of Figure 1 below.

**Figure 2:**
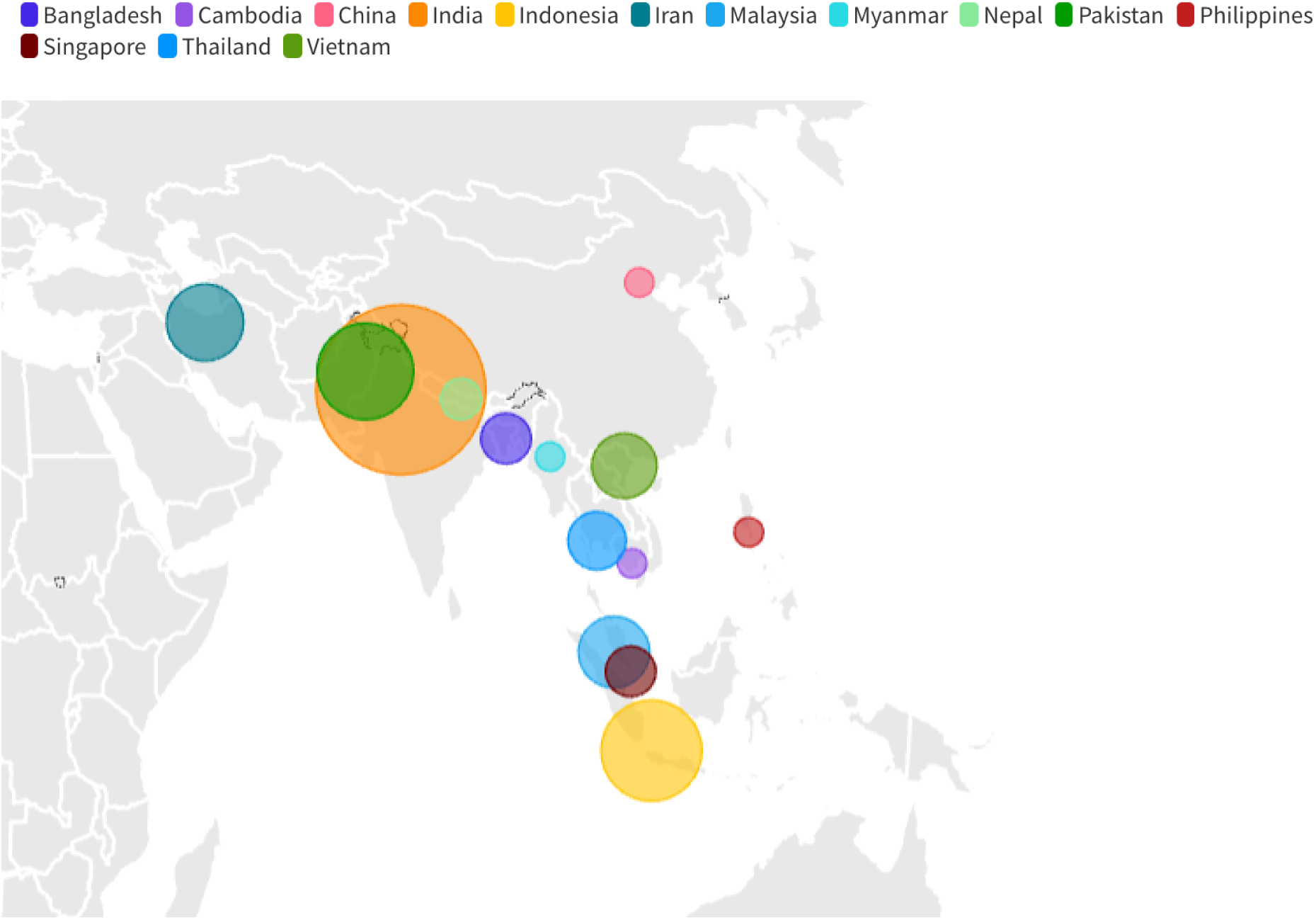
Countries included. Studies took place in India (34), Indonesia (12), Pakistan (10), Iran (7), Malaysia (6), Vietnam (5), Thailand (4), Bangladesh (3), Nepal (2), Singapore (3), Cambodia (1), China (1), Myanmar (1) and Philippines (1)

### Year of publication

When disaggregated by year of publication, we noticed that from 2019 onwards, the number of studies published significantly grew compared to years prior. It could be hypothesised that the increased number of publications in those years is linked to the overall rapid growth of published scholarly work due to relaxed and fast-tracked publication practices during the COVID-19 pandemic (23). However, that wouldn’t necessarily account for the higher number of publications in 2019, during pre-pandemic times. Tuberculosis (TB) was the primary disease target of eight out of the 12 studies published in 2019, suggesting a disease-specific focus from organisations or governments in that timeframe. This, however, would need to be further explored for more conclusive proof. A complete number of publications by year can be seen in Figure 3 below.

**Figure 3:**
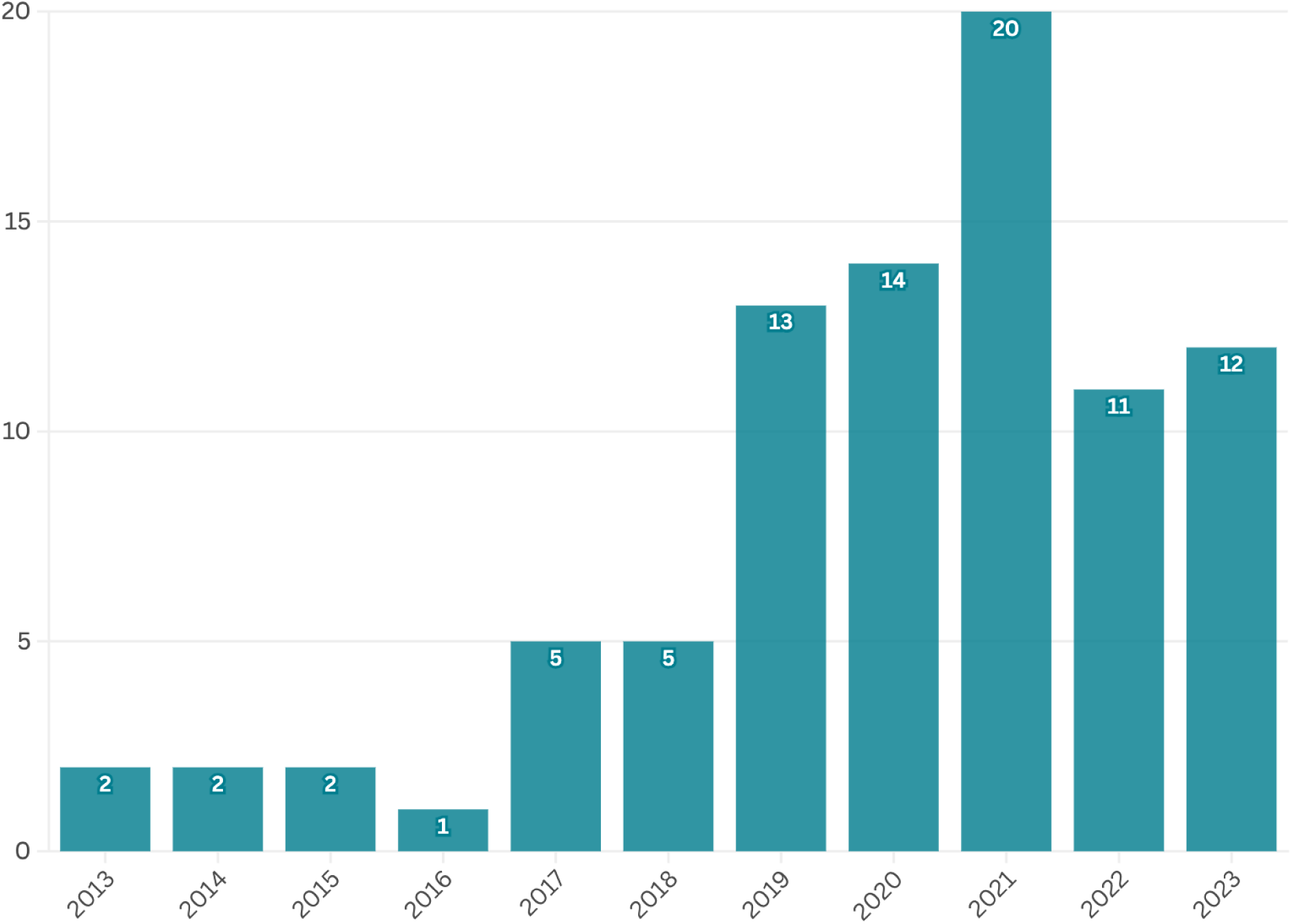
Number of studies published per year.

### Health concern

The disease/health concern on which researchers focused their attention the most was TB, with 52 studies. This was followed by smoking cessation interventions (12), air quality and air pollution interventions (9), asthma (8), chronic obstructive pulmonary disease (COPD) (2), and pneumonia (2). One of the study’s health focus was general lung disease (Figure 4). The high prevalence of studies investigating digital health interventions for TB may suggest that there is high interest, which translates into funding from all stakeholders (government, research institutions, iNGOs) in finding more effective ways to diagnose and treat TB in these regions (and likely in similar contexts across the world) via the use of digital health interventions.

**Figure 4:**
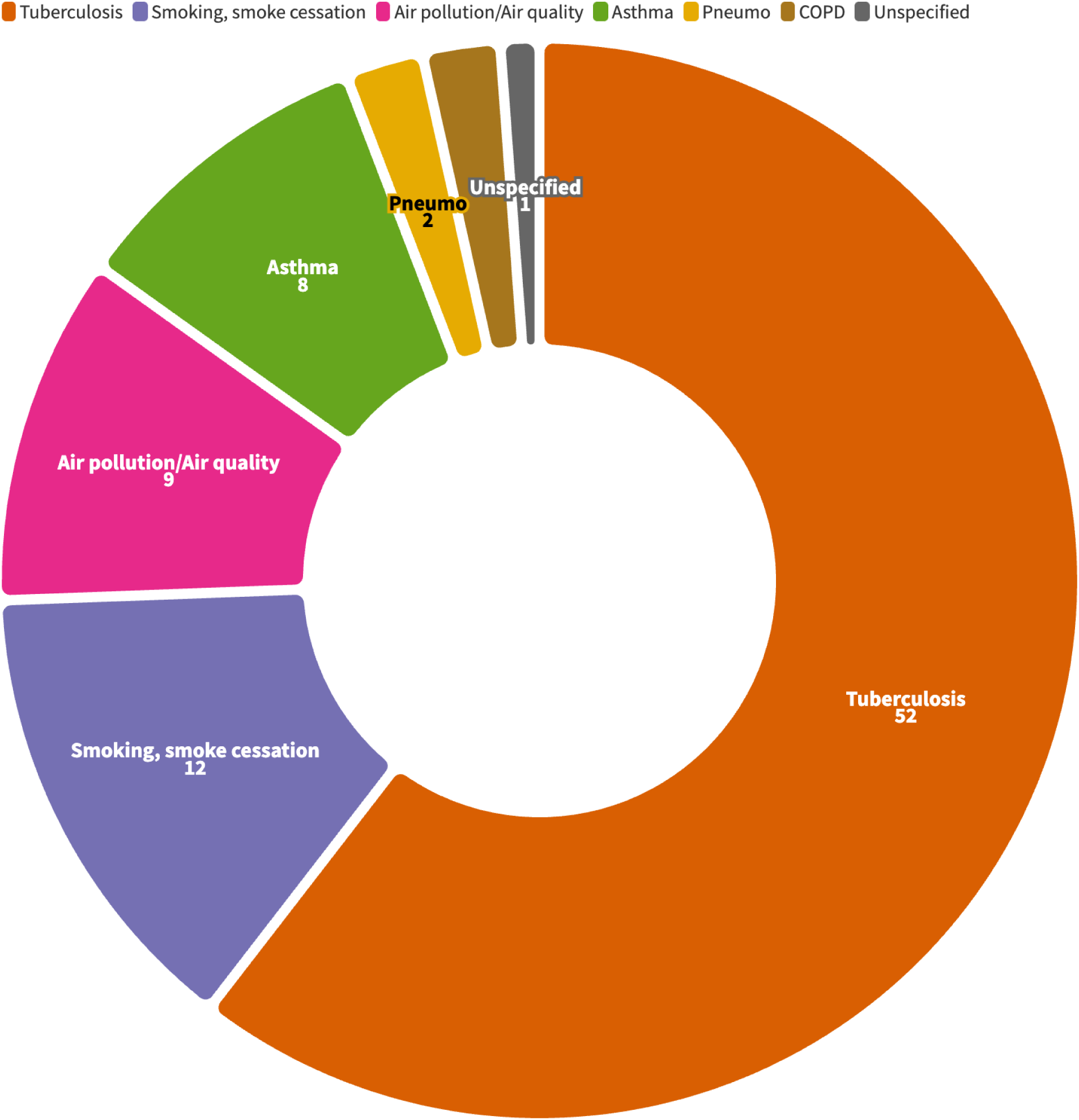
Disease/health concern

### Prototypes

Of 86 studies, 71 were pilot projects (Figure 5). Pilot projects can be defined as specific technologies deployed in particular settings for a limited time to assess their effectiveness, safety and cost-saving potential (24). Pilots are valuable and necessary when creating and deploying DHIs; however, the ‘pilotitis’ of the field has been a longstanding problem (8,25,26) whereby there are significant amounts of funding and energy devoted to creating and deploying proof-of-concept digital health tools without tangible plans to scale. This has resulted in a shortage of evidence of the efficacy and efficiency of digital health tools at scale, which hinders evidence-based decision-making for researchers, practitioners and policymakers in digital health matters. However, there were two different types of pilot projects in this study. First, the studies that developed a prototype DHI from scratch, that is, they created a dedicated piece of technology to carry out the study, with no further intention of upscaling beyond the study (or at least no such intention was expressed in the study manuscript). Second, the studies that, although they were pilot projects as defined above, used already existing and, many times, well-documented technological tools. These were primarily studies using SMS, phone calls, CAD4TB, 99DOTS, and ArcGIS.

**Figure 5:**
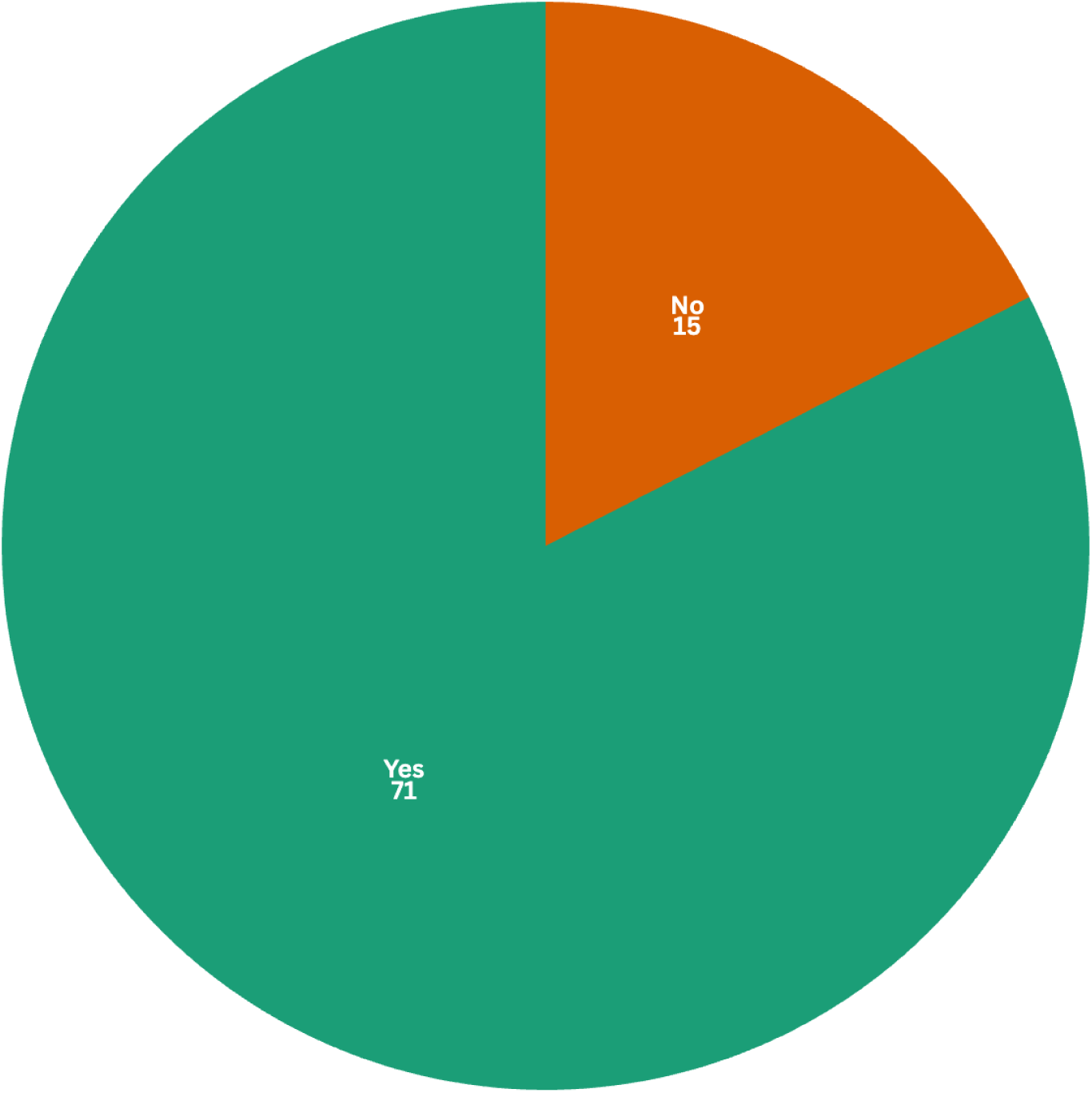
Interventions that were prototypes vs not

### WHO Type of digital health intervention

The WHO’s Classification of Digital Health Interventions ‘categorizes the different ways in which digital and mobile technologies are being used to support individuals and health system needs. It acknowledges the diverse ways in which digital capabilities are leveraged in different digital applications and services, to address personal and health system challenges and needs.’ (27) The four main categories in this classification are DHIs for Persons, DHIs for Healthcare Providers, DHIs for Health Managers and Support Personnel, and DHIs for Data Services. These, in turn, are subdivided into categories (Figure 6).

**Figure 6:**
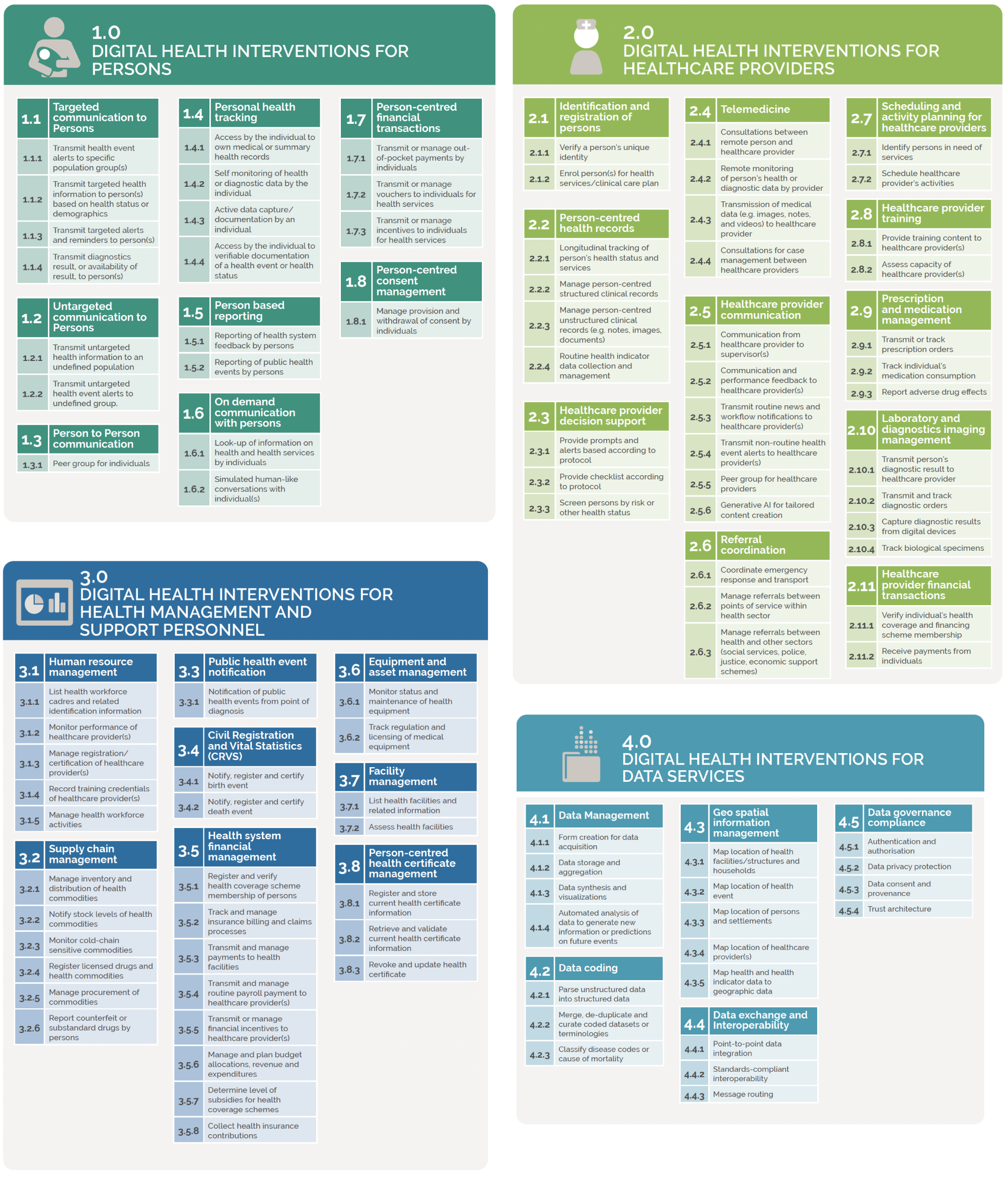
WHO Classification of DHIs categories and subcategories (27). (Used with permission under a CC License)

When looking at the studies included in this scoping review, we can see that DHIs for persons are the ones most widely used, followed by DHIs for data services. After that, DHIs for healthcare providers, managers, and support personnel are employed less in the studies included (Figure 7).

**Figure 7:**
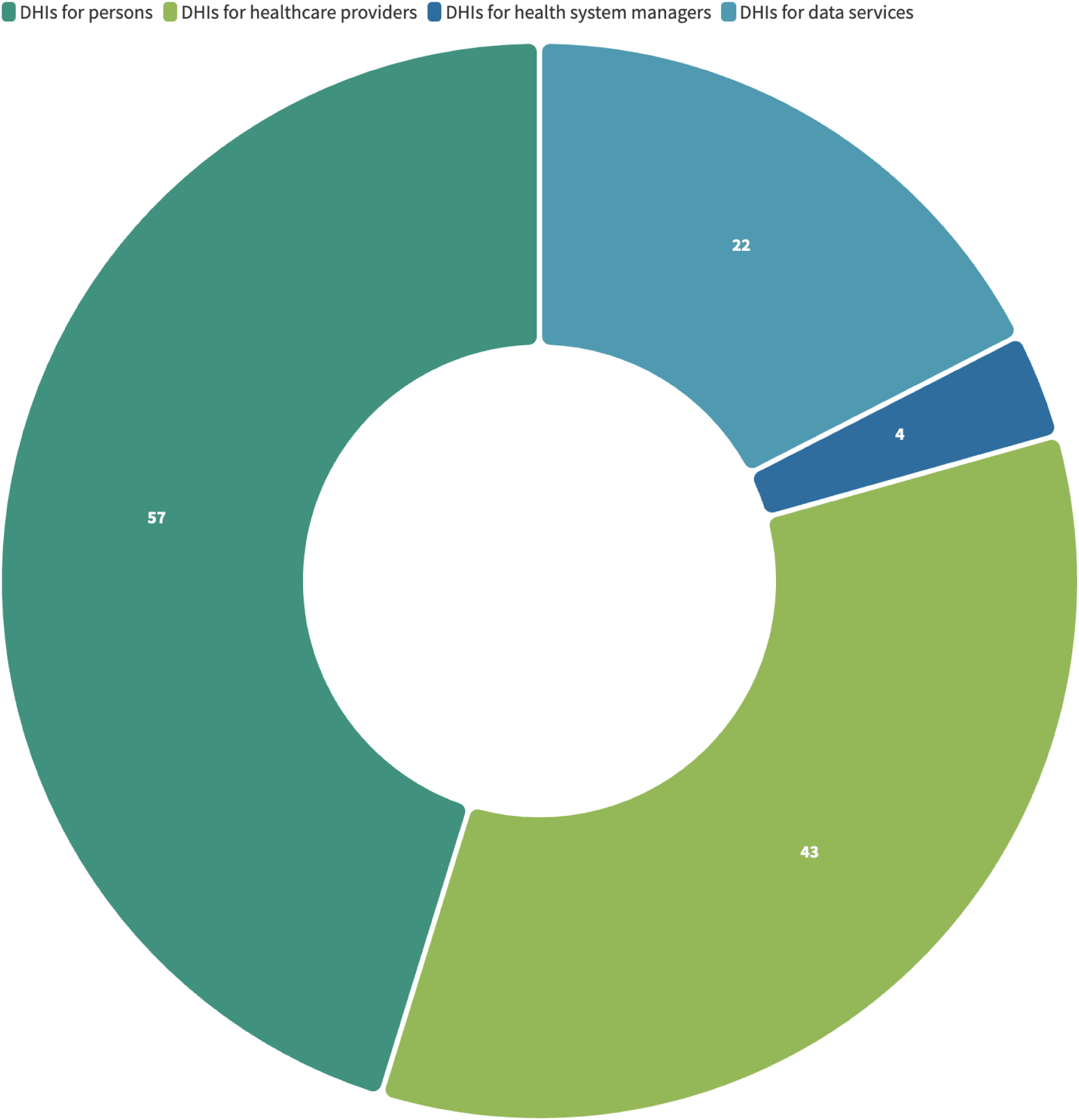
DHIs included in this study are classified according to the WHO’s Classification of DHIs categories.

When disaggregated by health concern/disease (Figure 8), we see that, for example, most of the interventions for smoking cessation are geared toward people and patients, which is also true of tools for asthma. Digital tools for air pollution and air quality monitoring and reporting primarily focus on data management and services, such as automated data analysis, to generate new information or predictions on future events.

**Figure 8:**
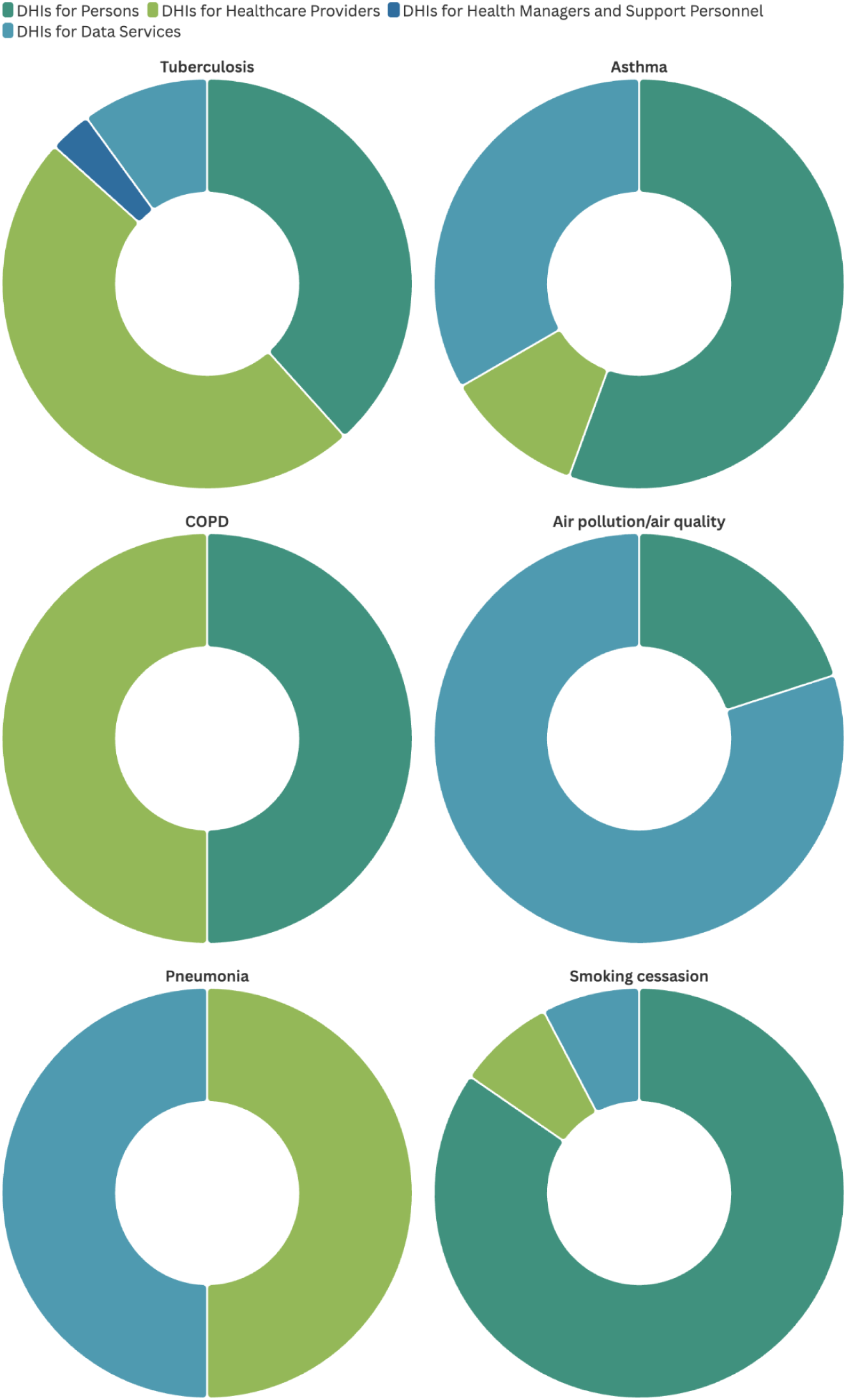
DHIs included in this study disaggregated by health concern.

### mERA checklist

The mHealth Evidence Reporting and Assessment (mERA) checklist aims to identify a minimum set of information needed to define a mHealth intervention (content), where it is being implemented (context), and how it was implemented (technical features), to support replication of the intervention (28). This standardisation checklist was chosen over others, such as CONSORT-EHEALTH (29), because of the mERA checklist’s global focus and the domains’ comprehensiveness. The aspects on which studies most frequently reported were the technology platform used, the way the intervention was delivered, the content of the intervention and how people were engaged and adopted the intervention (Figure 9). The least reported aspects were some of the more critical ones, such as interoperability, cost assessment, data security and regulation. The lack of reporting on these aspects likely reflects that most of the tools are pilot projects; therefore, there is no long-term approach to them. While understandable, this misses the opportunity to showcase a comprehensive view of these tools and establish their effectiveness and viability in the long term more firmly.

**Figure 9:**
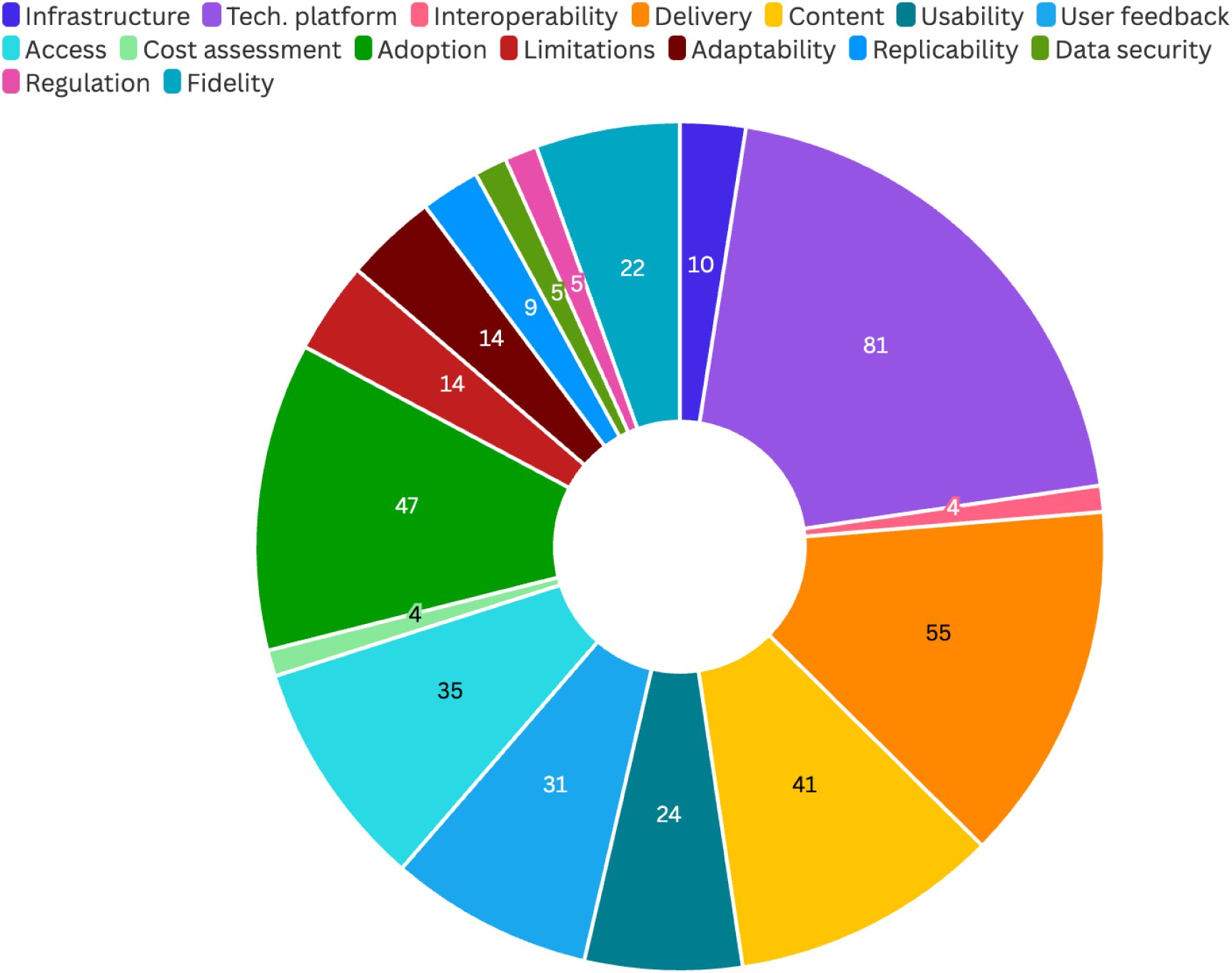
mERA fields and the reported frequency in the included studies

### Old technologies reimagined

When examining the types of technologies used in the studies, we can find a wide range, including older technologies, such as short message service (SMS) and phone calls. These approaches are still valid and effective in many settings due to their low cost and familiarity with the technology (30). However, some of the studies (31,32) showed a growing trend in digital health: using AI-powered chatbots (33). Those studies used these kinds of chatbots specifically for capacity building for health workers in the prevention and management of TB. Other studies (34,35) used what can be considered a new iteration of the SMS: WhatsApp. They used WhatsApp to transmit targeted messaging to patients the same way it would be done through an SMS.

Another old technology being ‘reimagined’ is the X-ray. Ten studies evaluated the accuracy of ML and AI technologies applied to chest X-rays (CXR) in patients with TB. Out of those, the majority studied the software CAD4TB (36–46). CAD4TB software can quickly interpret a CXR image using deep-learning technology and is designed for patients older than four. It is already being used in various settings with good performance (47). All of the studies found applying AI and ML to CXR interpretation proper and efficient, particularly in the countries included in this study. These countries have low economic resources, which impact the availability of Genexpert (the gold standard for TB diagnosis), and limited human resources, which affects the possibility of quickly reading CXRs by qualified radiologists.

### Artificial intelligence and machine learning

Beyond being used to screen patients by risk or health status (such as in the case of CAD4TB), AI and ML were employed for automated data analysis to generate new information or predictions on future events related to air quality and air pollution, tobacco use, TB and pneumonia (48–62). Although these studies found AI and ML beneficial and highly accurate, all were cautious about endorsing AI and LM at a large scale, recommending more extensive trials and research before scaling up.

The studies in this review show that AI and ML can have safe and effective real-life applications when integrated into clinical decision-support (CDDS) tools (49,61,63–65).

They can also be used as public health tools for predicting and forecasting adverse health effects such as increased air pollution or disease surveillance tools for infectious diseases such as TB or pneumonia.

### Tuberculosis

Of all the included studies, those that targeted TB represented 60% (n=52). This reflects the reality of the region since Southeast Asia has the highest incidence of TB in the world at 46% (66). As seen in Figure 7, digital tools are being used in all four categories of DHIs: for persons, for providers, for managers and support personnel and for data.

However, the two types of tools most commonly developed and used are tools to communicate with patients (SMS, phone calls, video calls) regarding their TB treatments and tools to aid clinicians in diagnosis via AI- and ML-powered CXRs interpretation, with 67 studies making use of tools within these two use cases. The results of the studies were favourable overall, showing that the technological tools employed fit the purpose and accomplish their stated objectives.

## Discussion

### Principal findings

Our review aimed to map the respiratory digital health interventions in South and Southeast Asia to identify existing technologies, opportunities, and gaps and to put forward pertinent recommendations based on the insights gained. This scoping review provides a snapshot (since technology may have already changed since we first ran our searches) of existing digital health tools in South and Southeast Asia being used for respiratory health. We have found that the number of DHIs employed matches the prevalence of diseases in the region, that most of the studies report on pilot projects and not mature interventions, and that AI and ML are becoming highly relevant players in digital health.

### Covid-19

Our scoping review protocol had COVID-19 as one of the diseases included in the search criteria. However, when screening started, we realised that including those studies was going to be challenging, therefore, we decided to exclude them. There were two main reasons for this. First, many studies involved other health issues such as inflammation, headaches and fatigue besides respiratory symptomatology or studies examined DHIs for long Covid, which consists of a constellation of health issues (67). It became time-consuming and difficult to screen for studies that only looked at respiratory symptoms related to COVID-19, and therefore, we were inclined to exclude all the studies that involved COVID-19. Second, the DHIs developed and deployed during COVID-19 had a very specific purpose and, like in previous large-scale outbreaks (68), were at risk of being abandoned or decommissioned once the pandemic ended, potentially not being used by the time of this review. These two factors drove the decision to exclude any study that involved COVID-19. We, however, acknowledge the missed learnings from this decision. Therefore, as future work, we believe that undertaking a scoping review similar to this one but only examining DHIs for COVID-19 respiratory health issues in South and Southeast Asia can benefit the scientific community.

### To prototype or not to prototype?

To answer our second research question (How are the current DHIs addressing (or not) the respiratory health needs of the region?), we must address the high prevalence of prototypes in our included studies. Almost 82% (n=71) of the studies involved a DHI that was a prototype or a pilot project. These studies reported, overall, favourable results on their project, and yet virtually none of them laid out concrete plans to scale and further test their DHI. No doubt, much insight can be gained from a pilot project, such as the appropriateness of the tool, the acceptability, usability, functionality, etc.; in fact, pilot projects are a necessary part of deploying DHIs. However, pilot projects, by definition, do not have an impact at the population level and, therefore, cannot address the respiratory issues faced by whole communities in South and Southeast Asia.

We offer two potential approaches to this issue so that progress can be made regarding DHIs meaningfully addressing the respiratory needs of whole communities. First, we need to focus on sustaining innovations. There are many definitions and classifications of innovations. Christensen et al. (69) talk about disruptive and sustaining innovations. Disruptive innovations are those that reinvent a technology or simply invent something new altogether while sustaining innovations seek to improve existing products and processes. Many of the pilot projects concentrated on creating a new piece of technology to address a specific respiratory health issue, which could be classified as an attempt to develop a disruptive innovation. However, focusing on repurposing, improving, or adapting existing technologies for specific contexts and respiratory health issues may be a better approach and one that has been used before (70). For example, some of the studies took already existing digital health tools and adapted them to their context or, as in the case of CAD4TB, evaluated them with local datasets to understand their performance. This can add significant value because it takes a well-established, well-functioning tool and evaluates it in a specific context. If the evaluation is favourable, the speed at which that tool, in this case CAD4TB, can be upscaled is much greater than a tool built from scratch because of its proven effectiveness and safety track record.

Yet, this is often not enough. Besides focusing more on studying and piloting sustaining innovations, a systematic and rigorous approach to pilot projects is paramount. Most of the included studies sought to evaluate narrow specifications (feasibility, usability, accuracy, reliability, etc.) instead of having a holistic perspective. When using the mERA checklist to understand what aspects of their DHIs authors reported on, it can be seen that several aspects are critically underreported. Failing to evaluate and report all the key factors that influence a DHI creates a distorted understanding of the potential an intervention has to scale and have a society-level impact. As we saw earlier, the characteristics of a prototype are their limited nature in terms of place (deployed in one place or just a few) and time (for a month, six months, or other limited amount of time). Beyond this, for a prototype to significantly fulfil its purpose, that is, its effectiveness, safety and cost-saving potential, it must encompass as many aspects of a real, large-scale implementation as possible. Using standardised checklists and toolkits such as mERA, a systematic procedure for developing, evaluating and reporting results can be applied. This, in turn, paints a more accurate picture of what it may entail to scale up a particular DHI, what resources may be needed, and what critical issues may arise on the way there.

### Nikshay and 99DOTS

At the other end of the spectrum from pilot projects, we can find Nikshay and 99DOTS. Nikshay “(Ni=End, Kshay=TB) is a web-based patient management system for TB control under the National Tuberculosis Elimination Programme (NTEP).” (71) and 99DOTS “is a low-cost approach for monitoring and improving TB medication adherence [that] enables remote observation of doses administered by patients or their family members.” (72) These two interventions are based in India (although 99DOTS is used elsewhere), and several of the studies in this review deal with different aspects of these two programs (73–80), evaluating the accuracy, acceptability, feasibility, data management and other aspects, but all within the context of these mature interventions.

99DOTS is now part of Nikshay, reflecting even greater sustainability as proven interventions merge to simplify work processes for patients and providers. There are some key things that can be learned from the Nikshay intervention. First, it proves true what is already known about the necessary pieces for a DHI to scale and be sustainable. Nikshay is a fitting example of the reality that institutionalisation in the form of government funding and support can have an impact at scale and that integration with other interventions through national data standardisation processes can simplify workflows (81,82). Moreover, it reflects the reality that flexible and ‘old’ technology that allows being deployed in resource-constrained and signal-poor areas can expand the areas in which a tool can be used and that conscientious capacity-building efforts that are built in the intervention ensures that all the healthcare workers that use these tools also have access to training. Finally, the Nikshay example shows how national-level policy is critical to support the development of DHIs (8,83,84).

The success of Nikshay does not imply that the intervention is perfect or that it does not face challenges. A report from Stop TB Partnerships highlights several areas of improvement for Nikshay, such as the need to adopt international interoperability standards, add real-time data functionality, eliminate duplicate, paper-based tasks, and expand infrastructure and capacity-building endeavours (84). Knowing the success, failures and areas of improvement of interventions such as Nikshay and 99DOTS over time, as they grow and scale, allows implementers and researchers to distil lessons learnt that can positively impact the development and deployment of future interventions. Moreover, the studies in this scoping review that look at the different aspects of Nikshay and 99DOTS are effectively sustaining innovations that seek to improve and expand upon a functioning and established intervention, potentially increasing their impact beyond what those studies’ authors could have done if they would have created something from the ground up.

### Artificial intelligence and machine learning

Interventions using AI and ML were abundant in this review and had highly positive accuracy results. However, the authors cautioned regarding endorsing such technologies, suggesting that more research is needed with larger datasets or in different settings.

The region of South and Southeast Asia can leapfrog by capitalising on AI and ML technologies early on, as they deploy DHIs that are AI/ML ready and capable instead of having to adapt legacy systems as other countries with a more mature digital health ecosystem (85). However, this competitive advantage needs to be used wisely. Some early best practices around AI are already emerging. These are predominantly focused on the ethical use of AI, preventing bias in AI and ML algorithms, considering health equity, utilising explainable AI (and not relying on the ‘black box’ explanation), stringent data privacy regulations and sufficiently large training datasets that match real-life data (86). Such practices and recommendations are to be carefully heeded so that the region can maximise the benefits of AI and ML while mitigating risks.

There is growing fear that AI and ML will erase healthcare jobs in the coming years, and although some administrative tasks may be automated in the future (87), AI and ML are not being developed to replace healthcare professionals. Instead, their primary purpose is to serve or enhance human skills and knowledge and facilitate faster, more accurate decision-making processes (88). The potential of AI and ML for air quality prediction, TB diagnosis, respiratory infectious disease incidence and transmission forecasting, and clinical decision support systems is hard to grasp fully. However, such potential will only be realised if ethical, equitable, and transparent policies, practices, and evaluation mechanisms for AI and ML are firmly established in the region.

### Recommendations

As a result of our findings and analysis, we propose several recommendations for the region regarding DHIs for respiratory health:

1. Assess the need for another pilot project: Thoroughly evaluate the need for a given pilot project and justify it. Consider whether building on what already exists is a better option for sustainability and impact.
2. Report on interventions systematically: Before starting the project, decide on the evaluation metrics and parameters used for reporting. Many existing frameworks can aid this process. In this scoping review, we have utilised mERA, but others can be used instead, depending on needs. Deciding beforehand will help gather metrics during the entire project. Such metrics and insights can be used for evaluation and reporting systematically and transparently. This increases the reliability of the project and allows the entire digital health community to learn more valuable lessons.
3. AI and ML are a growing field in the region with many valuable applications for respiratory health. To obtain the maximum benefit from them, emerging best practices must be followed so that interventions can be safe, effective, ethical, and equitable.

### Limitations

Several limitations exist in this scoping review. As previously mentioned, it is likely that between the time the databases were searched and the time of this writing (about a year difference), other publications exist presenting different DHIs for respiratory health due to the fast pace of technology development. Likewise, although this scoping review has searched peer-reviewed and grey databases, hundreds of other interventions likely exist in South and Southeast Asia, as a quick internet search can reveal.

Notwithstanding these limitations, the results and insights of this review are still valid. Including more DHIs may have yielded additional results, but it would not have made the results of this review invalid.

The specific geographical focus of this review made it impossible to include many studies that, although they may have information about some of the relevant countries for this study, were global in nature and did not disaggregate results by country. Insights from those studies were now analysed, potentially missing an opportunity to reinforce the findings of this review or uncover new ones.

As already mentioned, all COVID-19 studies were ultimately excluded due to their high number and lack of exclusive respiratory health focus. A COVID-19-only review could be undertaken to distil findings regarding DHIs for COVID-19 and understand whether those interventions are still in use and why.

The mERA checklist was originally developed to report on mobile-based DHIs. In this scoping review, we have taken the liberty of applying it to all the interventions included in this study because it was deemed to be a tool fit for the purpose of this review, which sought to understand the completeness of DHI reporting. No study used any predefined tool for evaluation and reporting; therefore, utilising the mERA checklist did not conflict with other reporting checklists employed by other authors.

## Conclusion

The burden of respiratory disease in South and Southeast Asia is staggering, severing years off people’s lives and preventing them from living healthy lives. RESPIRE2 seeks to highlight evidence-based interventions that can be adopted and adapted for long-term sustainability. This scoping review represents one such effort by collating and synthesising information and knowledge in the current state of DHIs in the region, showcasing how such interventions are or are not alleviating the respiratory health burden of disease and making relevant recommendations to researchers and practitioners. This scoping review helps create a base of knowledge from which disrupting or sustaining innovations can be forged so that they are planned, deployed, scaled and evaluated to have long-lasting positive impacts on the health and lives of the people we seek to serve.

## Author’s Contributions

Jay Evans and Laura Evans conceptualised the idea and screened the papers. Laura Evans guided the analysis and wrote the first draft. Jay Evans provided ongoing input and analysis. Jay Evans, Adina Abdullah, and Zakiuddin Ahmed provided feedback and recommendations for further manuscript drafts.

## Acknowledgements

Marshall Dozier, the librarian at the University of Edinburgh, provided feedback on the protocol’s search strategy and overall design.

## Data Availability

The data resulting from this scoping is available as supplementary material as well as the PRISMA-ScR checklist.

## References

1. Mahya FZ. A Brief History of Digital Health [Internet]. That Medic Network. 2021 [cited 2023 Mar 15]. Available from: https://medium.com/that-medic-network/a-brief-history-of-digital-health-b238f1f5883

2. Maaß L, Pan CC, Freye M. Mapping Digital Public Health Interventions Among Existing Digital Technologies and Internet-Based Interventions to Maintain and Improve Population Health in Practice: Protocol for a Scoping Review. JMIR Res Protoc [Internet]. 2022 Mar 31 [cited 2023 Mar 15];11(3):e33404. Available from: https://www.ncbi.nlm.nih.gov/pmc/articles/PMC9015775/

3. Roser M, Ritchie H, Spooner F. Burden of disease. Our World Data [Internet]. 2021 Sep 25 [cited 2023 Mar 15]; Available from: https://ourworldindata.org/burden-of-disease

4. Vos T, Lim SS, Abbafati C, Abbas KM, Abbasi M, Abbasifard M, et al. Global burden of 369 diseases and injuries in 204 countries and territories, 1990–2019: a systematic analysis for the Global Burden of Disease Study 2019. The Lancet [Internet]. 2020 Oct 17 [cited 2023 Mar 15];396(10258):1204–22. Available from: https://www.thelancet.com/journals/lancet/article/PIIS0140-6736(20)30925-9/fulltext#supplementaryMaterial

5. Soriano JB, Kendrick PJ, Paulson KR, Gupta V, Abrams EM, Adedoyin RA, et al. Prevalence and attributable health burden of chronic respiratory diseases, 1990–2017: a systematic analysis for the Global Burden of Disease Study 2017. Lancet Respir Med [Internet]. 2020 Jun 1 [cited 2023 Mar 15];8(6):585–96. Available from: https://www.thelancet.com/journals/lanres/article/PIIS2213-2600(20)30105-3/fulltext

6. Sheikh A, Campbell H, Balharry D, Baqui AH, Bogaert D, Cresswell K, et al. RESPIRE: The National Institute for Health Research’s (NIHR) Global Respiratory Health Unit. J Glob Health [Internet]. 2018 Dec [cited 2023 Mar 15];8(2):020101. Available from: http://jogh.org/documents/issue201802/jogh-08-020101.pdf

7. Hui CY, Abdulla A, Ahmed Z, Goel H, Monsur Habib GM, Teck Hock T, et al. Mapping national information and communication technology (ICT) infrastructure to the requirements of potential digital health interventions in low- and middle-income countries. J Glob Health [Internet]. [cited 2023 Mar 15];12:04094. Available from: https://www.ncbi.nlm.nih.gov/pmc/articles/PMC9804211/

8. Evans J, Bhatt S, Sharma R. The Path to Scale: Navigating Design, Policy, and Infrastructure. In: mHealth Innovation in Asia [Internet]. Springer, Dordrecht; 2018 [cited 2018 Mar 5]. p. 31–48. (Mobile Communication in Asia: Local Insights, Global Implications). Available from: https://link-springer-com.ezproxy.is.ed.ac.uk/chapter/10.1007/978-94-024-1251-2_3

9. Cresswell K, Williams R, Sheikh A. Developing and Applying a Formative Evaluation Framework for Health Information Technology Implementations: Qualitative Investigation. J Med Internet Res [Internet]. 2020 Jun 10 [cited 2023 Mar 15];22(6):e15068. Available from: https://www.jmir.org/2020/6/e15068

10. World Bank. Protocol: An evidence gap map for digital health interventions. [Internet]. [cited 2023 Mar 15]. Available from: https://documents1.worldbank.org/curated/en/211341591931421760/pdf/Study-Protocol-An-Evidence-Gap-Map-for-Digital-Health-Interventions.pdf

11. NIHR RESPIRE. IPCRG. [cited 2023 Mar 16]. NIHR RESPIRE. Available from: https://www.ipcrg.org/projects/research/nihr-respire

12. University of Edinburgh. The University of Edinburgh. 2022 [cited 2023 Mar 16]. What is RESPIRE? Available from: https://www.ed.ac.uk/usher/respire/about/what-is-respire

13. Arksey H, O’Malley L. Scoping studies: towards a methodological framework. Int J Soc Res Methodol [Internet]. 2005 Feb 1 [cited 2020 Aug 25];8(1):19–32. Available from: 10.1080/1364557032000119616

14. Peters M, Godfrey C, McInerney P, Munn Z, Trico A, Khalil H. Chapter 11: Scoping Reviews. In: Aromataris E, Munn Z, editors. JBI Manual for Evidence Synthesis [Internet]. JBI; 2020 [cited 2020 Aug 31]. Available from: https://wiki.jbi.global/display/MANUAL/Chapter+11%3A+Scoping+reviews

15. Evans L, Evans J, Fletcher M, Abdullah A, Ahmed Z. Mapping Respiratory Health Digital Interventions in South and Southeast Asia: Protocol for a Scoping Review. JMIR Res Protoc [Internet]. 2024 Jan 12 [cited 2024 Nov 5];13(1):e52517. Available from: https://www.researchprotocols.org/2024/1/e52517

16. The Joanna Briggs Institute. The Joanna Briggs Institute Reviewers’ Manual 2015: Methodology for JBI Scoping Reviews [Internet]. 2015 [cited 2020 Aug 25]. Available from: https://nursing.lsuhsc.edu/JBI/docs/ReviewersManuals/Scoping-.pdf

17. Evans L, Evans J, Pagliari C, Källander K. Exploring the Equity Impact of Current Digital Health Design Practices: Protocol for a Scoping Review. JMIR Res Protoc [Internet]. 2022 May 17 [cited 2022 Nov 10];11(5):e34013. Available from: https://www.researchprotocols.org/2022/5/e34013

18. Tricco AC, Lillie E, Zarin W, O’Brien KK, Colquhoun H, Levac D, et al. PRISMA Extension for Scoping Reviews (PRISMA-ScR): Checklist and Explanation. Ann Intern Med [Internet]. 2018 Sep 4 [cited 2021 May 27];169(7):467–73. Available from: https://www.acpjournals.org/doi/full/10.7326/M18-0850

19. UNSD — Methodology [Internet]. [cited 2023 Jul 5]. Available from: https://unstats.un.org/unsd/methodology/m49/

20. Covidence. Covidence. 2021 [cited 2021 Feb 24]. Covidence - Better systematic review management. Available from: https://www.covidence.org/

21. Suri H. Ethical Considerations of Conducting Systematic Reviews in Educational Research. In: Zawacki-Richter O, Kerres M, Bedenlier S, Bond M, Buntins K, editors. Systematic Reviews in Educational Research: Methodology, Perspectives and Application [Internet]. Wiesbaden: Springer Fachmedien; 2020 [cited 2020 Sep 16]. p. 41–54. Available from: 10.1007/978-3-658-27602-7_3

22. Grammarly. Grammarly [Internet]. 2024 [cited 2024 Nov 5]. Available from: https://grammarly.com/

23. Ioannidis JPA, Salholz-Hillel M, Boyack KW, Baas J. The rapid, massive growth of COVID-19 authors in the scientific literature. R Soc Open Sci [Internet]. 2021 Sep 7 [cited 2024 Nov 5];8(9):210389. Available from: https://royalsocietypublishing.org/doi/10.1098/rsos.210389

24. NHS. AI and Digital Regulations Service for health and social care. 2024 [cited 2024 Oct 9]. Piloting digital technologies in a health or care service. Available from: https://digitalregulations.innovation.nhs.uk/regulations-and-guidance-for-adopters/all-adopters-guidance/piloting-digital-technologies-in-a-health-or-care-service/

25. Kirkup B. Scaling up Health Service Delivery – From pilot innovations to policies and programmes. Public Health [Internet]. 2009 Sep [cited 2018 Mar 21];123(9):638–9. Available from: http://linkinghub.elsevier.com/retrieve/pii/S0033350609001541

26. Tomlinson M, Rotheram-Borus MJ, Swartz L, Tsai AC. Scaling Up mHealth: Where Is the Evidence? PLOS Med [Internet]. 2013 Feb 12 [cited 2018 Mar 5];10(2):e1001382. Available from: http://journals.plos.org/plosmedicine/article?id=10.1371/journal.pmed.1001382

27. World Health Organization. Classification of digital interventions, services and applications in health: a shared language to describe the uses of digital technology for health, 2nd ed [Internet]. 2023 [cited 2024 Jul 18]. Available from: https://www.who.int/publications/i/item/9789240081949

28. Agarwal S, LeFevre AE, Lee J, L’Engle K, Mehl G, Sinha C, et al. Guidelines for reporting of health interventions using mobile phones: mobile health (mHealth) evidence reporting and assessment (mERA) checklist. BMJ [Internet]. 2016 Mar 17 [cited 2022 Oct 11];352:i1174. Available from: https://www.bmj.com/content/352/bmj.i1174

29. Eysenbach G, Group CE. CONSORT-EHEALTH: Improving and Standardizing Evaluation Reports of Web-based and Mobile Health Interventions. J Med Internet Res [Internet]. 2011 Dec 31 [cited 2024 Nov 4];13(4):e1923. Available from: https://www.jmir.org/2011/4/e126

30. Suffoletto B. Deceptively Simple yet Profoundly Impactful: Text Messaging Interventions to Support Health. J Med Internet Res [Internet]. 2024 Aug 27 [cited 2024 Nov 27];26(1):e58726. Available from: https://www.jmir.org/2024/1/e58726

31. Modi B, Puwar B. Ni-kshay SETU, a Digital Health Intervention for Capacity Building in Tuberculosis under the National TB Elimination Program in India: A comprehensive mHealth app review. Healthline [Internet]. 2023 Dec 31 [cited 2024 Nov 4];14(4):342–7. Available from: https://www.healthlinejournal.org/index_pdf/507.pdf

32. Shah H, Patel J, Yasobant S, Saxena D, Saha S, Sinha A, et al. Capacity Building, Knowledge Enhancement, and Consultative Processes for Development of a Digital Tool (Ni-kshay SETU) to Support the Management of Patients with Tuberculosis: Exploratory Qualitative Study. J Med Internet Res [Internet]. 2023 Jun 19 [cited 2024 Nov 4];25(1):e45400. Available from: https://www.jmir.org/2023/1/e45400

33. Clark M, Bailey S. Chatbots in Health Care: Connecting Patients to Information: Emerging Health Technologies [Internet]. Ottawa (ON): Canadian Agency for Drugs and Technologies in Health; 2024 [cited 2024 Nov 4]. (CADTH Horizon Scans). Available from: http://www.ncbi.nlm.nih.gov/books/NBK602381/

34. Selvaraju S, Malaisamy M, Dolla CK, Murali L, Karikalan N, Saravanan B, et al. Application of mobile phone technology as intervention for the management of tuberculosis patients diagnosed through community survey. Indian J Med Res. 2022 Feb;155(2):301–5.

35. Ernirita E, Fahrudin A, Widiastuti E. The Effect of Social Media-based Pokemon Education on Adolescent Knowledge about Tuberculosis Prevention. Open Access Maced J Med Sci. 2022 Jun 12;10:992–7.

36. Geric C., Majidulla A., Tavaziva G., Nazish A., Saeed S., Benedetti A., et al. Artificial intelligence-reported chest X-ray findings of culture-confirmed pulmonary tuberculosis in people with and without diabetes. J Clin Tuberc Mycobact Dis [Internet]. 2023;31((Geric, Tavaziva, Benedetti, Ahmad Khan) McGill International TB Centre, Research Institute of the McGill University Health Centre, Montreal, Canada(Geric, Tavaziva, Benedetti, Ahmad Khan) Respiratory Epidemiology and Clinical Research Unit, Centre for Ou):100365. Available from: http://www.journals.elsevier.com/journal-of-clinical-tuberculosis-and-other-mycobacterial-diseases

37. Habib SS, Rafiq S, Zaidi SMA, Ferrand RA, Creswell J, Van Ginneken B, et al. Evaluation of computer aided detection of tuberculosis on chest radiography among people with diabetes in Karachi Pakistan. Sci Rep. 2020;10(1):6276.

38. Herman B., Sirichokchatchawan W., Pongpanich S., Nantasenamat C. Development and performance of CUHASROBUST application for pulmonary rifampicin-resistance tuberculosis screening in Indonesia. PLoS ONE [Internet]. 2021;16(3 March):e0249243. Available from: https://journals.plos.org/plosone/article/file?id=10.1371/journal.pone.0249243&type=printable

39. Herman B, Sirichokchatchawan W, Nantasenamat C, Pongpanich S. Artificial intelligence in overcoming rifampicin resistant-screening challenges in Indonesia: a qualitative study on the user experience of CUHAS-ROBUST. J Health Res. 2022;36(6):1018–27.

40. Murphy K, Habib SS, Zaidi SMA, Khowaja S, Khan A, Melendez J, et al. Computer aided detection of tuberculosis on chest radiographs: An evaluation of the CAD4TB v6 system. Sci Rep. 2020;10(1):5492.

41. Nash M, Kadavigere R, Andrade J, Sukumar CA, Chawla K, Shenoy VP, et al. Deep learning, computer-aided radiography reading for tuberculosis: a diagnostic accuracy study from a tertiary hospital in India. Sci Rep. 2020;10(1):210.

42. Nsengiyumva N.P., Hussain H., Oxlade O., Majidulla A., Nazish A., Khan A.J., et al. Triage of Persons With Tuberculosis Symptoms Using Artificial Intelligence-Based Chest Radiograph Interpretation: A Cost-Effectiveness Analysis. Open Forum Infect Dis [Internet]. 2021;8(12):ofab567. Available from: https://academic.oup.com/ofid

43. Qin Z.Z., Sander M.S., Rai B., Titahong C.N., Sudrungrot S., Laah S.N., et al. Using artificial intelligence to read chest radiographs for tuberculosis detection: A multi-site evaluation of the diagnostic accuracy of three deep learning systems. Sci Rep. 2019;9(1):15000.

44. Rahman MT, Codlin AJ, Rahman MM, Nahar A, Reja M, Islam T, et al. An evaluation of automated chest radiography reading software for tuberculosis screening among public- and private-sector patients. Eur Respir J. 2017;49(5).

45. SweetyBakyarani E., Srimathi H., Arul Leena Rose P.J. A comparative study on performance of pre-trained convolutional neural networks in tuberculosis detection. Eur J Mol Clin Med [Internet]. 2020;7(3):4852–8. Available from: https://ejmcm.com/article_5173_8a369f7be76eef8a84b2ad37d95afc11.pdf

46. Zaidi SMA, Habib SS, Van Ginneken B, Ferrand RA, Creswell J, Khowaja S, et al. Evaluation of the diagnostic accuracy of Computer-Aided Detection of tuberculosis on Chest radiography among private sector patients in Pakistan. Sci Rep. 2018;8(1):12339.

47. Murphy K, Habib SS, Zaidi SMA, Khowaja S, Khan A, Melendez J, et al. Computer aided detection of tuberculosis on chest radiographs: An evaluation of the CAD4TB v6 system. Sci Rep [Internet]. 2020 Mar 26 [cited 2024 Nov 4];10(1):5492. Available from: https://www.nature.com/articles/s41598-020-62148-y

48. Delavar M, Gholami A, Shiran G, Rashidi Y, Nakhaeizadeh G, Fedra K, et al. A Novel Method for Improving Air Pollution Prediction Based on Machine Learning Approaches: A Case Study Applied to the Capital City of Tehran. ISPRS Int J GEO-Inf. 2019;8(2).

49. Haider NS, Singh BK, Periyasamy R, Behera AK. Respiratory Sound Based Classification of Chronic Obstructive Pulmonary Disease: a Risk Stratification Approach in Machine Learning Paradigm. J Med Syst [Internet]. 2019;43(8):N.PAG-N.PAG. Available from: https://search.ebscohost.com/login.aspx?direct=true&db=jlh&AN=137490047&site=ehost-live

50. Hasan M, Faruk M, Biki B, Riajuliislatn M, Alam K, et al. Prediction of Pneumonia Disease of Newborn Baby Based on Statistical Analysis of Maternal Condition Using Machine Learning Approach. In 2021. p. 919–24.

51. Kumbalaparambi T.S., Menon R., Radhakrishnan V.P., Nair V.P. Assessment of urban air quality from Twitter communication using self-attention network and a multilayer classification model. Environ Sci Pollut Res Int. 2023;30(4):10414–25.

52. Rakholia R., Le Q., Quoc Ho B., Vu K., Simon Carbajo R. Multi-output machine learning model for regional air pollution forecasting in Ho Chi Minh City, Vietnam. Environ Int [Internet]. 2023;173((Rakholia, Le, Simon Carbajo) Ireland’s National Centre for Applied Artificial Intelligence (CeADAR), University College Dublin, NexusUCD, Belfield Office Park, Dublin, Ireland(Quoc Ho, Vu) Institute for Environment and Resources (IER), Ho Chi Minh City 7):107848. Available from: https://www.elsevier.com/locate/envint

53. Razavi-Termeh S.V., Sadeghi-Niaraki A., Choi S.-M. Asthma-prone areas modeling using a machine learning model. Sci Rep. 2021;11(1):1912.

54. Razavi-Termeh S, Sadeghi-Niaraki A, Choi S. Spatial Modeling of Asthma-Prone Areas Using Remote Sensing and Ensemble Machine Learning Algorithms. REMOTE Sens. 2021;13(16).

55. Sarkar N, Gupta R, Keserwani PK, Govil MC. Air Quality Index prediction using an effective hybrid deep learning model. Environ Pollut Barking Essex 1987. 2022;315(dvl, 8804476):120404.

56. Singh K, Meitei A, Alee N, Kriina M, Haobijam N. Machine learning algorithms for predicting smokeless tobacco status among women in Northeastern States, India. Int J Syst Assur Eng Manag. 2022;13(5):2629–39.

57. Tella A., Balogun A.-L. GIS-based air quality modelling: spatial prediction of PM10 for Selangor State, Malaysia using machine learning algorithms. Environ Sci Pollut Res Int. 2022;29(57):86109–25.

58. Ullah R., Khan S., Ali H., Chaudhary I.I., Bilal M., Ahmad I. A comparative study of machine learning classifiers for risk prediction of asthma disease. Photodiagnosis Photodyn Ther [Internet]. 2019;28((Ullah, Ali, Bilal) Agri. & biophotonics Division, National Institute of Lasers&Optronics, Islamabad, Pakistan(Khan) Department of Physics, Islamia College Peshawar, Pakistan(Chaudhary) Department of Bioinformatics and Biotechnology, International Islam):292–6. Available from: http://www.elsevier.com/wps/find/journaldescription.cws_home/701993/description#description

59. Usmani R.S.A., Pillai T.R., Hashem I.A.T., Marjani M., Shaharudin R., Latif M.T. Air pollution and cardiorespiratory hospitalization, predictive modeling, and analysis using artificial intelligence techniques. Environ Sci Pollut Res Int. 2021;28(40):56759–71.

60. Usmani R, Pillai T, Hashem I, Marjani M, Shaharudin R, Latif M. Artificial intelligence techniques for predicting cardiorespiratory mortality caused by air pollution. Int J Environ Sci Technol. 2023;20(3):2623–34.

61. Yellapu G.D., Rudraraju G., Sripada N.R., Mamidgi B., Jalukuru C., Firmal P., et al. Development and clinical validation of Swaasa AI platform for screening and prioritization of pulmonary TB. Sci Rep. 2023;13(1):4740.

62. Neo E, Hasikin K, Lai K, Mokhtar M, Azizan M, Hizaddin H, et al. Artificial intelligence-assisted air quality monitoring for smart city management. PEERJ Comput Sci. 2023;9.

63. Kaur R, Sharma A. An Accurate Integrated System to detect Pulmonary and Extra Pulmonary Tuberculosis using Machine Learning Algorithms. Intel Artif-IBEROAMERICAL J Artif Intell. 2021;24(68):104–22.

64. Mulyani E, Febriani S, Wiyono R, Ismanto R, Suciyono N, et al. Expert System For Comparative Diagnosis Probability Of Lung Disease Using Nearest Neighbour Algorithm. In 2021. p. 251–6.

65. Saybani M.R., Shamshirband S., Hormozi S.G., Wah T.Y., Aghabozorgi S., Pourhoseingholi M.A., et al. Diagnosing tuberculosis with a novel support vector machine-based artificial immune recognition system. Iran Red Crescent Med J [Internet]. 2015;17(4):e24557. Available from: http://ircmj.com/43768.pdf

66. WHO. Global Tuberculosis Report [Internet]. 2023 [cited 2024 Nov 4]. Available from: https://www.who.int/teams/global-tuberculosis-programme/tb-reports/global-tuberculosis-report-2023/tb-disease-burden/1-1-tb-incidence

67. NHS. nhs.uk. 2023 [cited 2024 Nov 5]. Long-term effects of COVID-19 (long COVID). Available from: https://www.nhs.uk/conditions/covid-19/long-term-effects-of-covid-19-long-covid/

68. PATH. Digital Square. 2022 [cited 2024 Nov 5]. COVID-19 Map & Match. Available from: https://digitalsquare.org/covid19-map-match

69. Christensen CM, Raynor ME, McDonald R. What Is Disruptive Innovation? Harvard Business Review [Internet]. 2015 Dec 1 [cited 2024 Nov 5]; Available from: https://hbr.org/2015/12/what-is-disruptive-innovation

70. Digital Square [Internet]. 2021 [cited 2024 Nov 28]. Partnering with countries to adapt digital health global goods for COVID-19 response. Available from: https://digitalsquare.org/blog/2021/8/10/partnering-with-countries-to-adapt-digital-health-global-goods-for-covid-19-response

71. Ministry of Health & Family Welfare, Government of India. Nikshay - About us [Internet]. 2024 [cited 2024 Nov 5]. Available from: https://nikshay.in/Home/AboutUs

72. 99DOTS. 99Dots. [cited 2024 Nov 5]. 99DOTS. Available from: https://99dots.org

73. Prabhu A, Agarwal U, Tripathy JP, Singla N, Sagili K, Thekkur P, et al. “99DOTS”techno-supervision for tuberculosis treatment – A boon or a bane? Exploring challenges in its implementation at a tertiary centre in Delhi, India. Indian J Tuberc [Internet]. 2020 Jan 1 [cited 2024 Nov 5];67(1):46–53. Available from: https://www.sciencedirect.com/science/article/pii/S0019570719303014

74. Thakkar D, Piparva KG, Lakkad SG. A pilot project: 99DOTS information communication technology-based approach for tuberculosis treatment in Rajkot district. Lung India Off Organ Indian Chest Soc. 2019;36(2):108–11.

75. Shah H, Patel J, Yasobant S, Saxena D, Saha S, Sinha A, et al. Capacity Building, Knowledge Enhancement, and Consultative Processes for Development of a Digital Tool (Ni-kshay SETU) to Support the Management of Patients with Tuberculosis: Exploratory Qualitative Study. J Med Internet Res. 2023 Jun 19;25:e45400.

76. Thomas BE, Kumar JV, Chiranjeevi M, Shah D, Khandewale A, Thiruvengadam K, et al. Evaluation of the Accuracy of 99DOTS, a Novel Cellphone-based Strategy for Monitoring Adherence to Tuberculosis Medications: Comparison of DigitalAdherence Data With Urine Isoniazid Testing. Clin Infect Dis Off Publ Infect Dis Soc Am. 2020 Dec 3;71(9):e513–6.

77. Thomas BE, Kumar JV, Onongaya C, Bhatt SN, Galivanche A, Periyasamy M, et al. Explaining Differences in the Acceptability of 99DOTS, a Cell Phone-Based Strategy for Monitoring Adherence to Tuberculosis Medications: Qualitative Study of Patients and Health Care Providers. JMIR MHealth UHealth. 2020 Jul 31;8(7):e16634.

78. Rodrigues R, Varghese SS, Mahrous M, Ananthaneni Kumar A, Ahmed MN, D’Souza G. Feasibility and acceptability pilot of video-based direct observed treatment (vDOT) for supporting antitubercular treatment in South India: a cohort study. BMJ Open. 2023 May 29;13(5):e065878.

79. Modi B, Puwar B. Ni-kshay SETU, a Digital Health Intervention for Capacity Building in Tuberculosis under the National TB Elimination Program in India: A comprehensive mHealth app review. Healthline [Internet]. 2023 Dec 31 [cited 2024 Nov 5];14(4):342–7. Available from: https://www.healthlinejournal.org/index_pdf/507.pdf

80. Hiwale M, Varadarajan V, Walambe R, Kotecha K. NikshayChain: A Blockchain-Based Proposal for Tuberculosis Data Management in India. Technologies [Internet]. 2023 Feb [cited 2024 Nov 5];11(1):5. Available from: https://www.mdpi.com/2227-7080/11/1/5

81. World Health Organization. World Health Organization. 2021 [cited 2023 Apr 11]. Global strategy on digital health 2020-2025. Available from: https://www.who.int/docs/default-source/documents/gs4dhdaa2a9f352b0445bafbc79ca799dce4d.pdf

82. PATH. The Journey to Scale: Moving Together Past Digital Health Pilots. 2014. Available from: https://www.path.org/resources/the-journey-to-scale-moving-together-past-digital-health-pilots/

83. Ministry of Health & Family Welfare, Government of India. Nikshay [Internet]. 2024 [cited 2024 Nov 5]. Available from: https://nikshay.in/

84. The Global Fund. Stop TB Partnership - India [Internet]. 2021 [cited 2024 Nov 5]. Available from: https://tbassessment.stoptb.org/India.html

85. Bhagat SV, Kanyal D. Navigating the Future: The Transformative Impact of Artificial Intelligence on Hospital Management- A Comprehensive Review. Cureus [Internet]. 2024 Feb 20 [cited 2024 Nov 5];16(2):e54518. Available from: https://pmc.ncbi.nlm.nih.gov/articles/PMC10955674/

86. Polevikov S. Advancing AI in healthcare: A comprehensive review of best practices. Clin Chim Acta [Internet]. 2023 Aug 1 [cited 2024 Nov 5];548:117519. Available from: https://www.sciencedirect.com/science/article/pii/S0009898123003212

87. Health Journal. Top 10 Healthcare Jobs that AI will Displace [Internet]. Health Journal. 2023 [cited 2024 Nov 5]. Available from: https://www.thehealthjournals.com/top-10-healthcare-jobs-that-ai-will-displace/

88. Sezgin E. Artificial intelligence in healthcare: Complementing, not replacing, doctors and healthcare providers. Digit Health [Internet]. 2023 Jul 2 [cited 2024 Nov 5];9:20552076231186520. Available from: https://pmc.ncbi.nlm.nih.gov/articles/PMC10328041/

